# Genetic susceptibility for major depressive disorder associates with trajectories of depressive symptoms across childhood and adolescence

**DOI:** 10.1101/2020.03.02.19007088

**Authors:** Alexandre A. Lussier, Matt Hawrilenko, Min-Jung Wang, Karmel W. Choi, Janine Cerutti, Yiwen Zhu, Major Depressive Disorder Working Group of the Psychiatric Genomics Consortium, Erin C. Dunn

**Author notes:** list in the Supplemental Materials.

## Abstract

**Background:** Early-onset depression during childhood and adolescence is associated with a worse course of illness and outcome than adult onset. However, the genetic factors that influence risk for early-onset depression remain mostly unknown. Using data collected over 13 years, we examined whether polygenic risk scores (PRS) that capture genetic risk for depression were associated with depression trajectories assessed from childhood to adolescence.

**Methods:** Data came from the Avon Longitudinal Study of Parents and Children, a prospective, longitudinal birth cohort (analytic sample=7,308 youth). We analyzed the relationship between genetic susceptibility to depression and three time-dependent measures of depressive symptoms trajectories spanning 4 to 16.5 years of age (class, onset, and cumulative burden). Trajectories were constructed using a growth mixture model with structured residuals. PRS were generated from the summary statistics of a genome-wide association study of depression risk using data from the Psychiatric Genomics Consortium, UK Biobank, and 23andme, Inc. We used MAGMA to identify gene-level associations with these measures.

**Results:** Youth were classified into 6 classes of depressive symptom trajectories: *high/renitent* (26.5% of youth), *high/reversing* (5.8%), *childhood decrease* (6.1%), *late childhood peak* (3%), *adolescent spike* (2.5%), and *minimal symptoms* (56.1%). PRS discriminated between youth in the late childhood peak, high/reversing, and high/renitent classes compared to the minimal symptoms and childhood decrease classes. No significant associations were detected at the gene level.

**Conclusions:** This study highlights differences in polygenic loading for depressive symptoms across childhood and adolescence, particularly among youths with high symptoms in early adolescence, regardless of age-independent patterns.

## INTRODUCTION

Major depressive disorder (MDD) is one of the most common, costly, and disabling mental disorders, with lifetime prevalence estimated at 11.7% among adolescents (Merikangas et al., 2010) and 16.6% among adults in the United States (Kessler et al., 2005). Whereas depression can emerge at any point in the life course, nearly one third of those who have depression report a first onset before age 21 (Zisook et al., 2007). These early-onset cases of depression, compared with later onset, have been associated with worse illness course and outcomes in adulthood, including increased risk for adult depression and later physical and mental health comorbidities (McLaughlin et al., 2012). However, depression is complex and developmentally heterogenous. Individuals with depression not only have different ages of first onset, but also show varying patterns of persistence once the disorder begins. Although prior cross-sectional studies have modeled this heterogeneity to identify more homogenous subgroups of depression, such studies are limited because depressive symptoms were assessed at a single point in time (Nandi, Beard, & Galea, 2009).

More recent longitudinal studies have sought to model this time-dependent heterogeneity in depressive symptoms (and internalizing symptoms more broadly) by classifying individuals into subgroups based on their symptom *trajectories* over time. These studies have prospectively and repeatedly assessed depressive symptoms across an average of 3 to 6 years of development (Ellis et al., 2017). Most studies have characterized between three and six primary trajectory classes, which include people with consistently low symptoms, chronically high symptoms, or decreases/increases in symptoms during childhood and adolescence. Importantly, this body of work has shown that subtyping depression by developmental trajectory can help identify the factors that shape the course of depressive symptoms over time.

Genetic mechanisms in particular may explain part of the heterogeneity in depressive symptoms across development (Cai, Choi, & Fried, 2020). Genome-wide association studies (GWAS) suggest that common genetic variation accounts for approximately 9% of the variance in the heritability of adult depression (Howard et al., 2019; Wray et al., 2018). Studies in children have also shown some links between genetic variation and depressive phenotypes, with a recent study finding associations between one genetic locus and depressive symptoms at age 13 in the ALSPAC cohort (Sallis et al., 2017). However, most GWAS have focused on presence versus absence of lifetime depression and/or cross-sectional measures, which do not account for depression heterogeneity (Lee et al., 2013). Thus, additional studies using longitudinal data are needed to enhance understanding of genetic predictors of depressive symptoms across time, especially because prior work has shown that estimates of depression heritability may change over time (Bergen, Gardner, & Kendler, 2007; Nivard et al., 2015).

In addition to longitudinal data, polygenic risk scores (PRS), which capture the additive effect of multiple alleles using summary statistics from GWAS, have been used recently to examine genetic liability to depression. PRS have been used to assess the aggregate impact of genetic contributions on depressive phenotypes at various ages. For example, PRS of depression have recently been associated with self- or maternal-reported childhood psychopathologies, such as internalizing symptoms from age 7 to 16 (Akingbuwa et al., 2020), emotional problem trajectories from age 4 to 17 (Riglin et al., 2018), clinically measured depressive symptoms between age 7 and 18 (Halldorsdottir et al., 2019), as well as self-reported adolescent depressive symptom trajectories from age 10 to 22 (Kwong et al., 2019; Rice et al., 2018). Although these studies have provided important new insights into the genetic underpinnings of depression, they collectively have 2 main limitations. First, most studies focus on narrower age ranges and few include children younger than 7 years old. Thus, there has been limited attention to the earliest manifestations of depressive symptoms, which is a shortcoming because some symptoms can emerge as early as preschool (Whalen, Sylvester, & Luby, 2017). Accounting for the complete age span when symptoms emerge and occur is needed to capture the full course of depressive symptoms during development and to identify the factors that drive and shape symptom trajectories across the life course. Second, because longitudinal studies have focused almost exclusively on age-related changes in symptoms, genetic links to age-independent patterns, such as changes in symptom length and recurrence frequency across time, are poorly understood. For example, young people may have characteristic patterns of responding to positive and/or negative life events that can influence the chronicity and recurrence of any given depressive episode (Hawrilenko, Masyn, Cerutti, & Dunn, 2020). Efforts to disentangle biological and environmental sources of this interindividual variability in depressive symptom patterns are needed.

To address these gaps and determine how polygenic factors shape depressive symptom trajectories, we examined the genetic contributions to depressive symptoms across a 13-year period (from age 4 to 16.5). To our knowledge, this is the longest time span examined for depressive (or internalizing) symptoms across childhood and adolescence. Recognizing that developmental heterogeneity encompasses not only symptom trajectories across time, but also the age at onset and burden of symptoms, we modeled the developmental patterns of depressive symptoms using three measures. First, we assessed classes of depressive symptom trajectories, constructed using a new form of growth mixture modeling that includes structured residuals to account for symptom changes over time (GMM-SR; Hawrilenko et al., 2020). Second, we modeled onset of depressive symptoms at age 4, coinciding with the earliest time period reported in prior studies (Whalen et al., 2017). Third, we examined the cumulative burden of depressive symptoms, reflecting the overall level of depressive symptoms across childhood and adolescence. We assessed both polygenic and gene-specific mechanisms on these three measures in two ways. We first tested whether our measures were associated with polygenic risk for depression, using summary statistics generated from recent large-scale GWAS of depression (Howard et al., 2019). We then performed the first genome-wide, gene-level analysis of longitudinal depressive phenotypes to determine whether specific genes were implicated in these associations.

## MATERIALS AND METHODS

### Cohort and analytic sample

Data came from the Avon Longitudinal Study of Parents and Children (ALSPAC), a large population-based birth cohort out of Avon, England of children followed from before birth through early adulthood (Boyd et al., 2013; Fraser et al., 2013) (**supplemental materials**). The current study analytic sample consisted of 7,308 children who met the following inclusion criteria: singleton births with genotype data and at least one measure of depressive symptoms completed between 4 and 16.5 years of age (**Figure S1**). Compared to the total original ALSPAC sample, youths in our analytic sample were more likely to be white, have slightly higher birth weights, and whose mothers were older, married, and had higher employment, higher education, and fewer previous pregnancies (**Table S1**).

### Genotyping, quality control, and imputation

9,912 youths in the full ALSPAC cohort were genotyped on the Illumina HumanHap550-Quad genotyping platform (Illumina Inc., San Diego, CA). After quality control, 500,527 directly genotyped SNPs and 8,082 youths remained, of whom 7,308 (90.4%) were included in our analyses, based on the exclusion criteria above (**Figure S1; supplemental materials**).

### Depressive symptom measures

Depressive symptom scores were derived from the Strengths and Difficulties Questionnaire (SDQ; Goodman, 1997) and the Short Mood and Feelings Questionnaire (SMFQ; Angold, Costello, Messer, & Pickles, 1995), both of which were completed by caregivers via mail. Briefly, depressive symptoms were measured with two of the five SDQ subscales, *child emotional problems* and *peer difficulties*, as well as with the SMFQ in adolescents. Using multiple measures helped ensure that the most developmentally relevant depressive symptoms were tapped at each age (Graham, Taylor, Olchowski, & Cumsille, 2006). The SDQ subscales captured “internalizing symptoms” at seven timepoints: ages 4, 7, 8, 9.5, 11.5, 13, and 16.5 (Goodman, 2001), while the SMFQ assessed depressive symptoms in adolescents at three timepoints: ages 11.5, 13, and 16.5 (Angold et al., 1995). We used confirmatory factor analysis to combine information across all questionnaire items into a single factor score representing the latent cause of their shared variability, which we interpret as *depressive symptoms* (**Table S2; supplemental materials**) (Hawrilenko et al., 2020; Kline, 2015). Full details of the measurement invariance analyses can be found in the technical supplement for Hawrilenko and colleagues (2020). Factor score estimates of this latent score were used in the subsequent growth mixture models (Curran et al., 2018).

### Depressive symptom trajectories

In a previous analysis, Hawrilenko and colleagues used a novel form of growth mixture modeling with structured residuals (GMM-SR) in MPlus (version 8.0) to classify youths into distinct subgroups using their patterns of change in depressive symptoms (Hawrilenko et al., 2020; Muthén & Muthén, 2017). The GMM-SR partitions individual variability in depressive symptoms over time into: (1) an average level, (2) age-dependent trajectories, and (3) group-specific, time-anchored patterns of change. The latter were modeled through the structured residual, which captures the difference between the observed depression score and the score predicted by the age-dependent trajectory at any timepoint. Thus, the residual represents the influence of unmodeled events, adding an autoregressive structure to how these unmodeled events relate over time. Positive values represent the degree to which a deviation from average symptom levels perpetuates across time (renitent responding), whereas negative values represent the tendency for symptoms to fluctuate above and below an average level (reversing responding). Importantly, the GMM-SR allowed us to analyze not only depressive symptom levels as a function of age (age-dependent patterns), but also how symptom levels respond to change from unmodeled life events (everything except age) within and between youths over time.

To characterize depressive symptoms across childhood and adolescence, we analyzed three time-dependent measures of depressive symptoms trajectories: (1) trajectory classes; (2) onset of symptoms; and (3) cumulative burden of symptoms. The methods used to construct these variables are described below.

### Depressive symptom trajectory classes

Based on work by Hawrilenko et al. (2020), we characterized six overarching classes of depressive symptom trajectories within our analytic sample (**Figure 1**): 1) *minimal symptoms* (stable and low symptoms across development), 2) *adolescent spike* (low childhood symptoms spiking to high levels in adolescence), 3) *late childhood peak* (symptoms increasing through middle childhood and decreasing over adolescence), 4) *childhood decrease* (high symptoms decreasing across childhood), 5) *high/reversing* (high symptoms with sharp oscillations between ages), and 6) *high/renitent* (high symptoms with few oscillations between ages). The model-estimated proportion of youths assigned to each trajectory class was 49.9% minimal symptoms, 2.5% adolescent spike, 3.3% late childhood peak (3%), 7.3% childhood decrease, 9.1% high/reversing, and 27.9% high/renitent.

**Figure 1.**
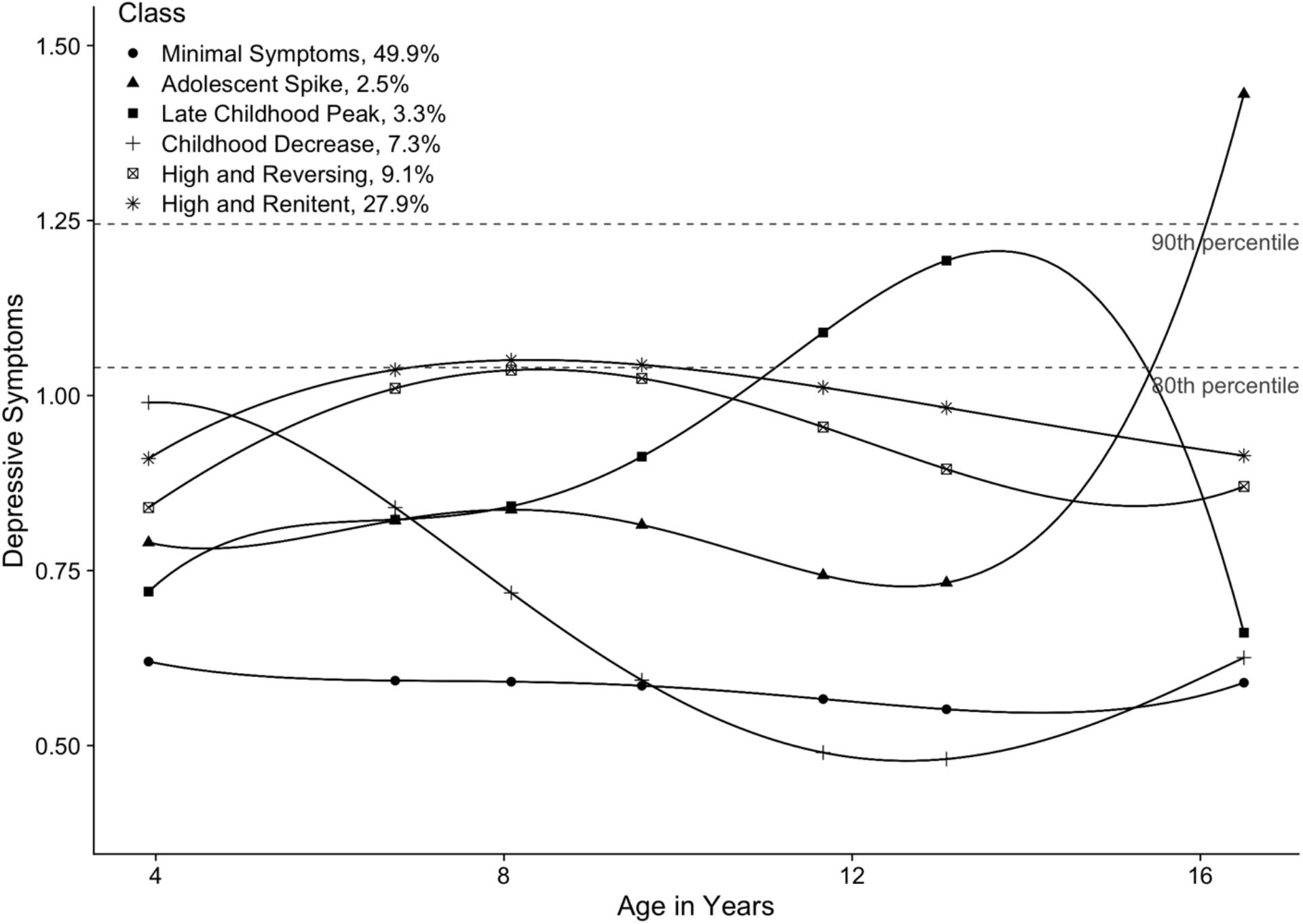
Classes of depressive symptom trajectories. Six classes were characterized using latent score analysis across ages 4 to 16.5 in 7,308 youths of the ALSPAC cohort. The latent scores are a composite score that captures depressive symptoms measured from the SDQ and SMFQ. Youth with higher scores have more depressive symptoms at that specific age. The percent of individuals assigned to each trajectory class is based on the model-estimated proportions. These are slightly different from the modal class proportions used in the MAGMA analysis, as they take into account the second and third place class assignments for each individual.

### Onset of depressive symptoms

Second, we measured the earliest possible onset of depressive symptoms, captured by the intercept of each youth’s depressive symptom trajectory at age 4. This intercept corresponded to baseline depressive symptoms latent score at age 4 for each individual (values ranged from 0.20 to 1.56, mean= 0.75; **Table S3**), where lower values correspond to less depressive symptoms. Given that clinically relevant depressive symptoms can emerge at preschool age (i.e., around age 4), early manifestations of depression may be more genetically driven and reflect stronger preexisting biological vulnerabilities than symptoms that emerge at later ages (Luby, 2010; Whalen et al., 2017).

### Cumulative burden of depressive symptoms

Third, we measured cumulative burden of depressive symptoms using the area under the curve (AUC) of the depressive symptom trajectory (Mills et al., 2018). We calculated the AUC as the integral of the function describing depressive symptom trajectories, estimated using individual growth factors (intercept, slope, quadratic, cubic and quartic terms). Thus, youths with higher depressive symptoms over developmental time display higher a cumulative burden of symptoms (AUC values ranged from 0.28 to 2.53, mean=1.13; **Table S3**). Of note, this measure was highly correlated with the onset of depressive symptoms (r=0.909), suggesting that the two measures likely tap similar dimensions of depressive symptom trajectories.

### Analyses

We performed our analysis in two main stages. All analyses were adjusted for sex and the top 10 principal components to account for population stratification.

### Polygenic risk score (PRS) analysis

We constructed PRS for depression using the latest summary statistics from the Psychiatric Genomics Consortium GWAS of MDD, wave 2 (PGC-MDD2; 43,204 cases; 95,680 controls), UK Biobank (127,552 cases; 233,763 controls), and 23andme (75,607 cases; 231,747 controls) in PLINK v.1.07 (Howard et al., 2019; Purcell et al., 2007) (**supplemental materials**). To assess polygenic influences on depressive symptom trajectory classes, we included PRS scores as predictors of class membership in MPlus, using the two-step method to account for classification uncertainty (Bakk & Kuha, 2018). The first step estimated the unconditional six-class GMM-SR in the current analytic sample. In the second step, the growth model parameters (e.g., intercepts, slope terms, autoregressive terms) were fixed at the estimates found in step 1, while class thresholds and covariate effects were freely estimated. We standardized PRS as z-scores so that all reported odds ratios (OR) could be interpreted as the effect of moving up or down one standard deviation of PRS on the odds of class membership. We used the Wald omnibus test to assess the relationship between PRS and depressive symptom trajectories. Multinomial logistic regressions were then used to contrast the model-estimated probability of assignment between different classes (Hawrilenko et al., 2020). Since the trajectories represent different groups of youths, these pairwise comparisons allowed us to understand the specific patterns of depressive symptoms differentiated by PRS. Parallel to the class-based analysis, we tested the relationship between PRS and depressive symptom onset or cumulative burden using linear regressions in R (version 3.6.1). We adjusted all PRS analyses for multiple comparisons using a false discovery rate (FDR) threshold of 5% (Benjamini & Hochberg, 1995).

### Gene-level analyses

To assess specific genetic influences on trajectory features, we performed genome-wide, gene-level associations with our three outcome measures using regression analyses in MAGMA (version 1.06)(de Leeuw, Mooij, Heskes, & Posthuma, 2015). For our gene-based analysis of trajectory classes, we focused on the two largest and most conceptually different classes, *minimal symptoms* and *high/renitent*. Because no tools currently exist to account for measurement error/classification uncertainty inherent to GMM in genome-wide analyses, we used modal class assignment to generate a case/control subset of youths in these two classes (**supplemental materials**). Although this approach may lead to liberal estimates of standard errors by not accounting for the uncertainty associated with GMM, point estimates remained unbiased (Vermunt, 2017). Briefly, each youth received a probability of assignment to each class, summing to 100%, and were assigned to the class in which they had the highest probability (i.e., modal class). The average probability of assigned class across all individuals was 76% (SD = 18%; **Figure S2**). Using this approach, 4,416 youths were classified as *minimal symptoms* (controls) and 1,895 youths were classified as *high/renitent* (cases). Sensitivity analyses using a higher probability threshold or continuous class probabilities for the high/renitent class can be found in the **supplemental materials**.

Our analyses of depressive symptom onset and cumulative burden used the continuous variables described earlier across all youths (N=7,308). A significance threshold of p<2.93e-6 was set to account for multiple testing of 17,044 genes; a nominal significance threshold was set at p<1e-4.

### Sensitivity analysis of maternal PRS

Given that depressed mothers may be more likely to rate their children as depressed, we performed a sensitivity analysis to determine if our results were biased by the use of maternal reports for youths’ depressive symptoms. To this end, we included maternal PRS for depression as predictors of child depressive symptom trajectories in 5,301 mother-child pairs (**supplemental materials**). Since mother and youth PRS were strongly correlated (r = 0.52), we expected a decrease in youth PRS effects when controlling for maternal PRS.

## RESULTS

### PRS were associated with depressive symptom trajectory classes

Our initial analysis revealed a significant relationship between PRS for depression and depressive symptom trajectories (45.8, p<1e-4; **Figure S3**), showing that the probability of class membership varied across PRS (**Figure 2**). To understand which specific patterns within trajectories drove these results, we examined pairwise contrasts between each pair of trajectories for a total of 15 contrasts (**Table 1; Figure S4**). Four contrasts showed statistically significant differences following multiple-test correction (FDR<0.05). Specifically, one standard deviation increase in PRS was associated with increased odds of assignment to the late childhood peak (OR=1.30, 95% CI: 1.10-1.54), high/reversing (OR=1.23, 95% CI: 1.07-1.41), or high/renitent classes (OR=1.28, 95% CI: 1.17-1.38), as compared to the minimal symptoms class. Similarly, each one-unit increase in the standardized PRS was associated with increased odds of being assigned to the high/renitent class compared to the childhood decrease class (OR=1.22, 95% CI: 1.05-1.42). When accounting for maternal PRS, these effects decreased an average of 34% (range: 28% to 45%) in statistically significant contrasts (**Table S4; supplemental materials**).

**Table 1.**
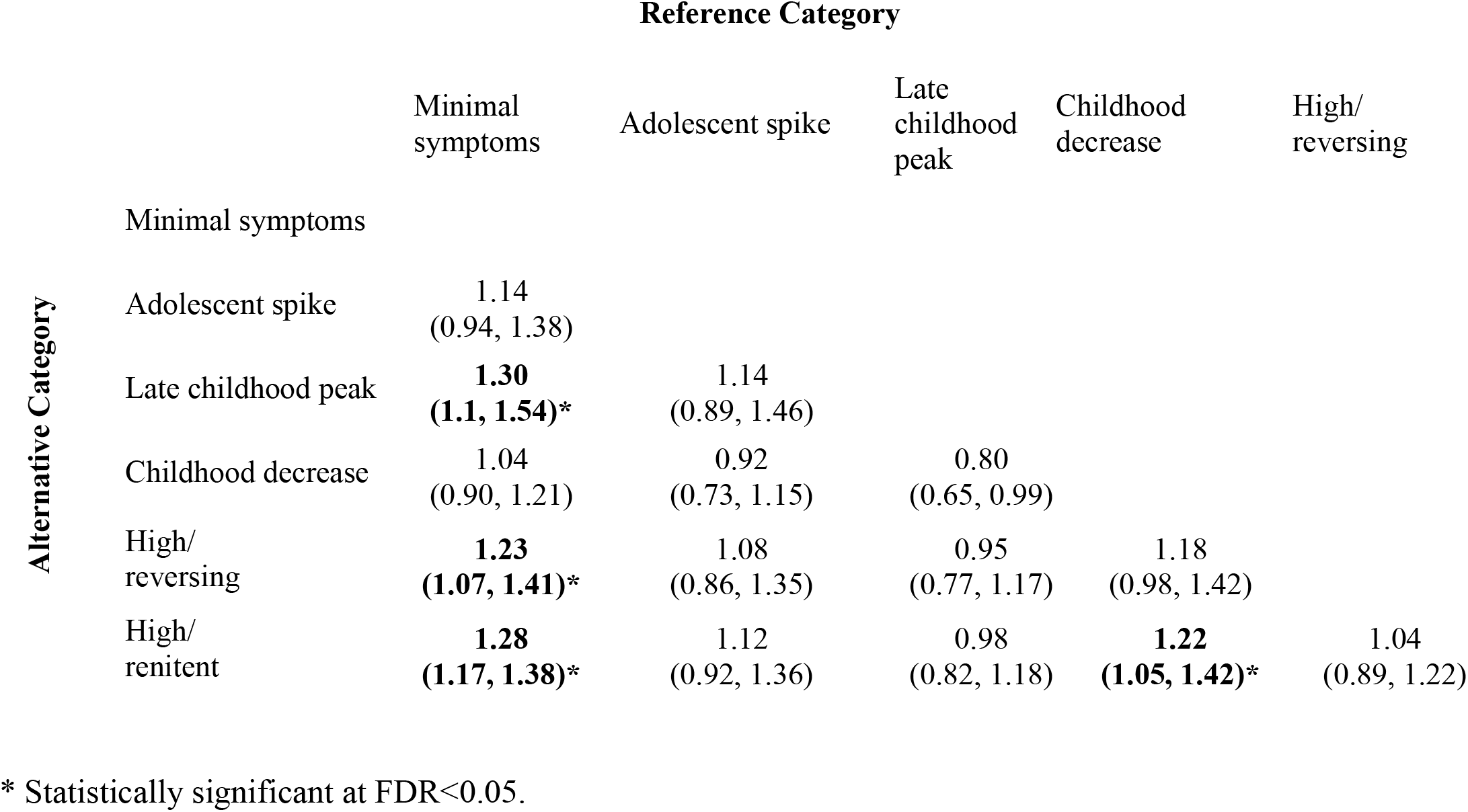
Odds of reference class assignment for each one standard deviation difference in standardized PRS.

|  |  | Reference Category |  |  |  |  |
| --- | --- | --- | --- | --- | --- | --- |
|  |  | Minimal symptoms | Adolescent spike | Late childhood peak | Childhood decrease | High/reversing |
| Alternative Category | Minimal symptoms |  |  |  |  |  |
|  | Adolescent spike | 1.14<br>(0.94, 1.38) |  |  |  |  |
|  | Late childhood peak | **1.30**<br>**(1.1, 1.54)\*** | 1.14<br>(0.89, 1.46) |  |  |  |
|  | Childhood decrease | 1.04<br>(0.90, 1.21) | 0.92<br>(0.73, 1.15) | 0.80<br>(0.65, 0.99) |  |  |
|  | High/reversing | **1.23**<br>**(1.07, 1.41)\*** | 1.08<br>(0.86, 1.35) | 0.95<br>(0.77, 1.17) | 1.18<br>(0.98, 1.42) |  |
|  | High/renitent | **1.28**<br>**(1.17, 1.38)\*** | 1.12<br>(0.92, 1.36) | 0.98<br>(0.82, 1.18) | **1.22**<br>**(1.05, 1.42)\*** | 1.04<br>(0.89, 1.22) |

\* Statistically significant at FDR<0.05.

**Figure 2.**
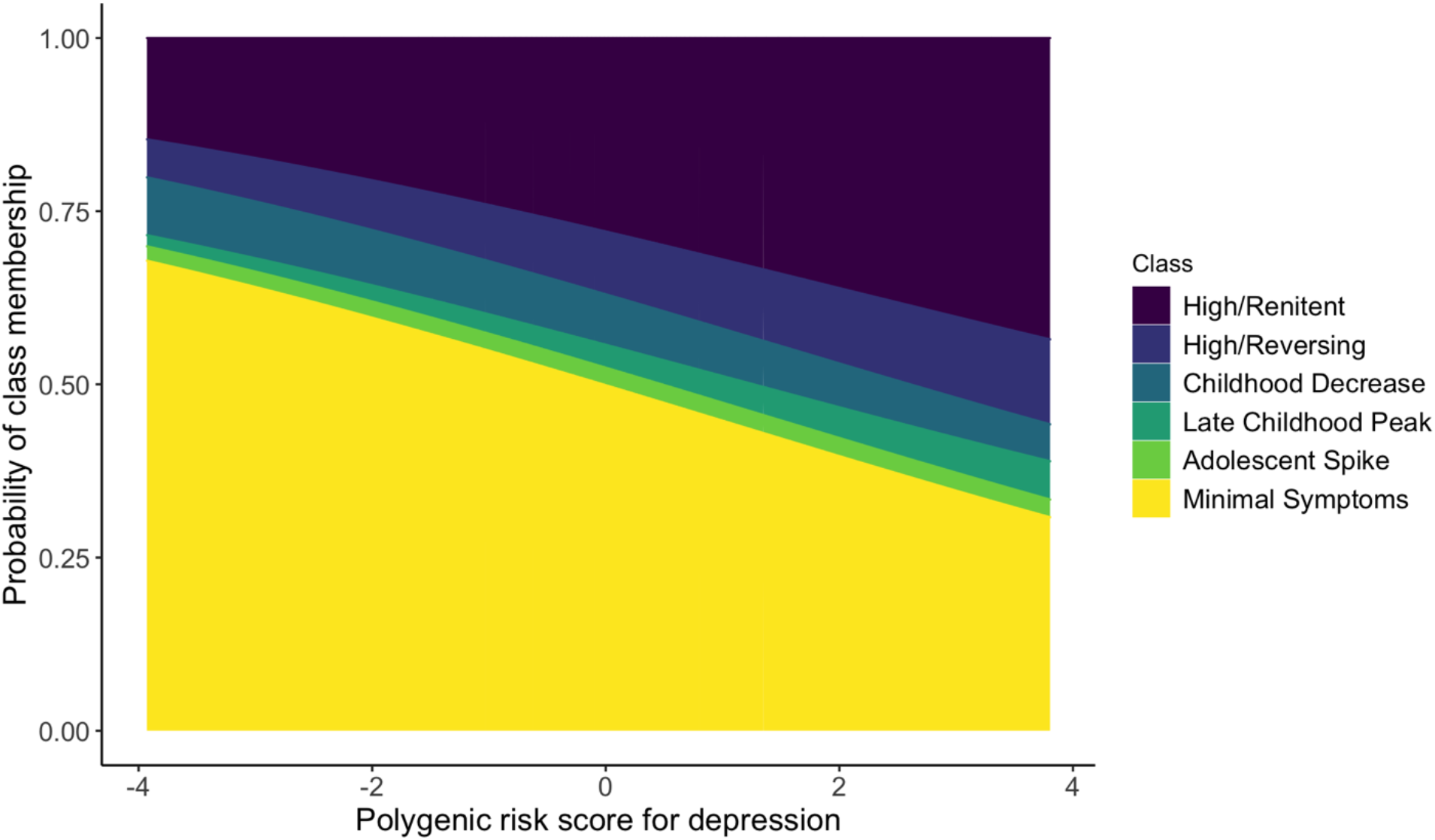
The probability of assignment to depressive symptom trajectory classes was skewed with PRS. The probability of class membership was calculated from the multinomial logistic regressions of estimated class probability. Negative PRS indicate lower genetic risk for depression whereas positive PRS indicate higher risk. All classes showed differences in probability of assignment based on PRS, except the adolescent spike class.

### PRS were associated with depressive symptom onset and cumulative burden

We next assessed whether PRS might influence features of depressive symptom trajectories beyond class alone. We found that higher PRS were associated with higher onset depressive symptoms (p=2.1e-10, β=0.018) and higher overall cumulative burden of depressive symptoms (p=4.1e-16, β =0.042) across all youths.

### Gene-based analyses revealed nominal associations with depressive symptom trajectory class, onset, and cumulative burden

Finally, we attempted to identify specific genes associated with our three outcome measures. No genes reached statistical significance at the genome-wide level (p<2.93e-6) when comparing youths in the high/renitent class to those in the minimal symptoms class, or when modeling onset and cumulative burden of depressive symptoms (**Figures S5, S6; Table S5**). However, seven different genes were nominally associated with depressive symptom features (GABRA4, LRR1, SIX5, DMPK, DKK1, SMDT1, DWMD). Of note, SIX5 and DMPK were nominally associated with both onset and cumulative burden of depressive symptoms.

## DISCUSSION

In this study, we examined the role of genetic contributions in shaping developmental patterns of depressive symptoms trajectories (class, onset, and cumulative burden) across a 13-year age range that spanned childhood and adolescence. Using the GMM-SR, we characterized six independent depressive symptom trajectories, which provided additional insight into the specific patterns of depressive symptoms that may be driven by genetic or environmental factors (Hawrilenko et al, 2020). Using trajectory classes, onset, and cumulative burden of depressive symptoms, we showed that polygenic influences may shape depressive symptom trajectories across development.

Similar to previous studies, our results showed that genetic liability may best discriminate between generally high versus low patterns of depressive symptoms during development (Kwong et al., 2019; Rice et al., 2018). However, our results extended beyond this known relationship, showing for the first time that these associations hold true irrespective of age-associated patterns of responding (i.e., renitent and reverse responding). As age-associated patterns of responding capture how youths respond to life events, fluctuations in depressive symptoms between timepoints may reflect coping mechanisms that are environmentally driven or learned, rather than genetic (Waugh & Koster, 2015).

Our results also further refined the time period when PRS may influence high depressive symptoms, showing that increased genetic risk for depression may manifest through elevated depressive symptoms during late childhood to early adolescence (approximately age 10-15). This result was in line with previous work, which shows that PRS of depression were associated with early-adolescent onset depressive symptom trajectories (Rice et al., 2018). As such, early adolescence may be a period when symptoms linked to genetic liability for depression are more likely to emerge.

Of note, the childhood decrease class has not been previously described in the depression genetics literature. This class was closer to the minimal symptoms class in terms of genetic influences, which may reflect maternal or other environmental influences. Indeed, Hawrilenko and colleagues (2020) reported that higher maternal psychopathology and education distinguish the childhood decrease class from the minimal symptoms class. The contrast between the childhood decrease and high/renitent classes also showed one of the largest decreases in effect size when maternal PRS for depression was included in the analysis (**supplemental materials**). Thus, the childhood decrease class may represent youths who are at lower risk for depression, but whose early-life depressive symptoms are driven by early life events not captured by the depression PRS. Since genetics are stable throughout the life course, it is possible that genetic vulnerability or resilience to depression could manifest throughout childhood and adolescence and that while depressive symptom trajectories vary, individual risk for depression later in life may remain the same. Previous work has shown that the impact of genetics on depression (i.e., heritability) increases over time, suggesting that some youths may self-select environments that reinforce their genetic susceptibility to depression (Bergen et al., 2007).

The only depressive symptom trajectory not associated with PRS was the adolescent spike class. Although the small size of this class may have affected our ability to detect significant associations, we detected associations between PRS and other classes of moderate size. As such, membership to this class may be primarily driven by environmental factors or sex-specific mechanisms, as shown by Hawrilenko and colleagues (2020). Indeed, the adolescent spike class showed an inflection point consistent with periods related to both internal (i.e., puberty) and external factors previously associated with depression, such as social environment, academic testing, and the like (Graber, Lewinsohn, Seeley, & Brooks-Gunn, 1997). However, these results are in contrast to previous work showing that depression PRS is associated with trajectories that arise later in adolescence or early adulthood (Kwong et al., 2019; Rice et al., 2018). These findings highlight the importance of multiple timepoints and broader developmental periods when assessing the factors influencing depressive symptoms, as shorter time periods may not have captured these distinct patterns of developmental heterogeneity. Future studies with access to data extending into adulthood may be able to further refine these trajectories and determine whether they do indeed reflect depressive symptom trajectories that continue into adulthood.

Beyond the polygenic influences of depression, we found no significant associations between individual genes and depressive symptom trajectories. Whereas the lack of associations may, in part, be due to small sample size, the genes identified at more relaxed p-value thresholds may reflect pathways impacting the manifestation of symptoms during development. For instance, GABRA4, a member of the GABAergic pathway, was nominally significant in the analyses of youths in the high/renitent and minimal symptoms classes. As this pathway has been previously associated with depression, our results could potentially point to a role in shaping developmental profiles of depression (Luscher, Shen, & Sahir, 2011). In contrast to our PRS results, our lack of significant gene-level associations may also emphasize polygenicity of depression, where its characteristics cannot be attributed to specific genes. Nevertheless, more comprehensive genome-wide analyses, such as gene-sets or pathway analyses, in larger samples may extend these findings by providing insight into the specific pathways shaping patterns of depressive symptoms across development.

Finally, previous work using these trajectories has shown that maternal psychopathology, abuse, and neighborhood-level disadvantage may increase the risk of belonging to all classes other than the minimal symptoms group (Hawrilenko et al., 2020). Given that depressive symptoms are highly correlated with environmental factors in this sample, the high variation in depressive symptom onset, cumulative burden, and overall trajectory may be attributed to factors not captured by genetics alone (Hawrilenko et al., 2020; Smoller et al., 2019). Genetic and environmental factors may also interact to drive depressive symptom trajectories, leading to the more complex phenotypes observed in our study.

## LIMITATIONS

There are some limitations to the present study. Aside from the relatively small sample size of this cohort for genetic analyses, this longitudinal sample is subject to attrition and self-selection over time and is composed of mainly European-ancestry youths, reducing the generalizability of our findings to other populations (Boyd et al., 2013). Depressive symptom scores were generated from maternal reports only, which may introduce inconsistencies over time or bias in reporting through residual maternal depression. However, when controlling for maternal depression PRS in our analyses, we only observed a small decrease in the effects of child PRS on depressive symptoms trajectories (**supplemental materials**). These results suggested that maternal polygenic risk was not as strong a predictor as the child’s own polygenic risk, despite child depressive symptoms being reported by their mothers. The PRS were also primarily generated from studies of depression in European adults, limiting the interpretability of findings in non-European populations (Howard et al., 2019). In addition, PRS built on summary statistics from adults may not be entirely reflective of genetic influences on depressive symptoms during early life. Nevertheless, as childhood and adult depression are highly correlated, our results likely capture a subset of youths that display increased genetic predisposition to lifetime depression. Finally, PRS still explain a very small portion of the variance in depression (1%-2%), which limits their ability to fully explain the genetic mechanisms influencing depressive symptom trajectories and may reduce their application in clinical settings (Martin, Daly, Robinson, Hyman, & Neale, 2019).

## CONCLUSIONS

Overall, our findings point to PRS as potential detectable early risk factors for depressive phenotypes during childhood and adolescence, particularly in youth with higher depressive symptoms during early adolescence. Findings provide new insights into how polygenic influences may shape depressive symptom trajectories. These findings may ultimately lead to the identification of genetic factors that shape age of depression onset and overall disease course, providing early targets to guide depression prevention efforts.

KEY POINTS
- The genetic factors that influence early-onset depression and depressive symptoms over developmental time remain mostly unknown.
- PRS for depression can discriminate between high and low patterns of depressive symptoms during development, particularly during early adolescence.
- PRS may be detectable risk factors for early-onset depressive phenotypes, particularly in youth with higher depressive symptoms.
- Further research is needed to understand the environmental and biological mechanisms driving depressive symptom trajectories.

## Data Availability

The data for the present study are available through the Avon Longitudinal Study of Parents and Children (ALSPAC).

http://www.bristol.ac.uk/alspac/

## ACKNOWLEDGEMENTS

This research was funded by the National Institute of Mental Health of the National Institutes of Health under Award Numbers K01MH102403 and R01MH113930 (Dunn), and T32MH017119 (Choi). The content is solely the responsibility of the authors and does not necessarily represent the official views of the National Institutes of Health. We are extremely grateful to all the families who took part in this study, the midwives for their help in recruiting them, and the whole ALSPAC team, which includes interviewers, computer and laboratory technicians, clerical workers, research scientists, volunteers, managers, receptionists and nurses. The UK Medical Research Council and the Wellcome Trust (Grant ref: 102215/2/13/2) and the University of Bristol provide core support for ALSPAC. A comprehensive list of grants funding is available on the ALSPAC website (http://www.bristol.ac.uk/alspac/external/documents/grant-acknowledgements.pdf). GWAS data were generated by Sample Logistics and Genotyping Facilities at Wellcome Sanger Institute and LabCorp (Laboratory Corporation of America) using support from 23andMe. We thank the research participants and employees of 23andMe for making this work possible. This publication is the work of the authors who will serve as guarantors for the contents of this paper. We thank Khalil Zlaoui and Susan Landry for their contributions and Dr. Kathryn Masyn for her input on the growth mixture models.

## ETHICAL CONSIDERATIONS

Ethical approval for the study was obtained from the ALSPAC Ethics and Law Committee and the Local Research Ethics Committee. Informed consent for the use of data collected via questionnaires and clinics was obtained from participants following the recommendations of the ALSPAC Ethics and Law Committee at the time.

## CORRESPONDENCE

- Erin C. Dunn, Psychiatric and Neurodevelopmental Genetics Unit, Center for Genomic Medicine, Massachusetts General Hospital, 185 Cambridge Street, Simches Research Building 6th Floor, Boston, MA 02114.
- Alexandre A. Lussier, Psychiatric and Neurodevelopmental Genetics Unit, Center for Genomic Medicine, Massachusetts General Hospital, 185 Cambridge Street, Simches Research Building, 5th Floor, Boston, MA 02114

**Abbreviated title:** Genetic contributions to depressive symptom trajectories

**Abbreviations**
ALSPAC: Avon Longitudinal Study of Parents and Children
GMM-SR: growth mixture model with structured residuals
MDD: major depressive disorder
PRS: polygenic risk scores

